# miRNA profiling in renal carcinoma suggest the existence of a group of pro-angionenic tumors in localized clear cell renal carcinoma

**DOI:** 10.1101/2019.12.12.19014696

**Authors:** Lucía Trilla-Fuertes, Natalia Miranda, Daniel Castellano, Rocío López-Vacas, Carlos A. Farfán Tello, Guillermo de Velasco, Felipe Villacampa, Elena López-Camacho, Guillermo Prado-Vázquez, Andrea Zapater-Moros, Enrique Espinosa, Juan Ángel Fresno Vara, Álvaro Pinto, Angelo Gámez-Pozo

**Author notes:** These authors contributed equally to this work (Lucía Trilla-Fuertes, Natalia Miranda). **Corresponding authors:** AP; AG-P.

## Abstract

Renal cell carcinoma comprises a variety of entities, the most common being the clear-cell, papillary and chromophobe subtypes. These subtypes are related to different clinical evolution; however, most therapies have been developed for clear-cell carcinoma and there is not a specific treatment based on different subtypes. In this study, one hundred and sixty-four paraffin samples from primary nephrectomies for localized tumors were analyzed. MiRNAs were isolated and measured by microRNA arrays. Significance Analysis of Microarrays and Consensus Cluster algorithm were used to characterize different renal subtypes. The analyses showed that chromophobe renal tumors are a homogeneous group characterized by an overexpression of miR 1229, miR 10a, miR 182, miR 1208, miR 222, miR 221, miR 891b, miR 629-5p and miR 221-5p. On the other hand, clear cell renal carcinomas presented two different groups inside this histological subtype, with differences in miRNAs that regulate focal adhesion, transcription, apoptosis and angiogenesis processes. Specifically, one of the defined groups had an overexpression of proangiogenic microRNAs miR185, miR126 and miR130a. In conclusion, differences in miRNA expression profiles between histological renal subtypes were established. In addition, clear cell renal carcinomas had different expression of proangiogenic miRNAs. With the emergence of antiangiogenic drugs, these differences could be used as therapeutic targets in the future or as a selection method for tailoring personalized treatments.

## Introduction

Renal carcinoma (RC) is the sixth most common cancer in men and the eight in women, with 73,820 estimated new cases and 14,770 estimated deaths in the United States in 2019 [1]. Two thirds of patients have localized disease and an additional 16% have locoregional disease (stage III) at diagnosis. A significant proportion of all these patients (up to 40% in stage III) will eventually relapse [2, 3].

Antiangiogenic multi-kinase inhibitors have demonstrated significant efficacy in the metastatic setting, but have not fulfilled expectations in the adjuvant setting. Sorafenib (SORCE trial), pazopanib (PROTECT trial) and axitinib (ATLAS trial) failed to improve disease-free survival when compared with placebo, whereas sunitinib improved disease-free survival but did not impact in overall survival (STRAC trial) [4-7]. As a consequence, sunitinib has been approved for adjuvant therapy by the Food and Drug Administration, but not by the European Medicines Agency and observation remains the standard of care after resection.

The current classical classification of renal carcinoma refers to subtypes that have been named on the basis of predominant cytoplasmic or architectural features, anatomic location, correlation with a specific disease background, as well as molecular alterations or familial syndromes [8]. The Cancer Genome Atlas has made considerable efforts to molecularly characterized different neoplasms, amongst them, renal carcinoma, establishing molecular characteristics of the different histological renal subtypes [9, 10]. So far, this information has not contributed to improve the personalized treatment for patients with renal cell carcinoma.

Molecular markers different from gene expression could improve our understanding of this disease. MicroRNAs are small RNA sequences which regulate different cellular processes [11]. They are good molecular biomarkers or even therapeutic targets, especially in clinical paraffin samples [12]. For these reasons, miRNAs have acquired importance as biomarkers in cancer.

The aim of this study is to determine miRNA profiles which allow us to characterize RC subtypes. Interestingly, we identified two groups of clear cell renal carcinoma (ccRCC) tumors, one of them with an overexpression of pro-angiogenic microRNAs.

## Material and methods

### Samples

One hundred and sixty-four patients diagnosed with localized RC were recruited for this study. An observational study was carried out, where all radical and partial nephrectomies performed at Hospital Universitario 12 de Octubre in Madrid between 1999 and 2008 were included. Written informed consent was obtained from all patients. The protocol was approved by the Ethical Committee of Hospital Universitario 12 de Octubre. The evolution of these patients was obtained from clinical records.

### miRNA isolation and quantification

396 miRNAs were measured from 164 renal formalin-fixed paraffin-embedded (FFPE) tumor samples. microRNA extraction and sample processing were done as previously described [13]. Briefly, selected FFPE tumor specimens were cut into serial sections with a thickness of 10 µm. Total RNA was then isolated using the miRNEasy Kit (Quiagen). Purified RNA quality control for quantity and purity was assessed using an ND-1000 NanoDrop spectrophotometer (Thermo Fisher Scientific).

### MicroRNA Arrays

Samples were hybridized to Human miRNA Microarray Release 14.0, 8×15K (Agilent Technologies). MicroRNA Labeling Kit (Agilent Technologies) was used to label RNA. Basically, 100 ng of total RNA were dephosphorylated and Cyanine 3-pCp molecule was ligated to the 3′ end of each RNA molecule by using T4 RNA ligase. One hundred ng of Cy3 labeled RNA were hybridized for 20 hours at 55°C in a hybridization oven (G2545A, Agilent) set to 20 rpm in a final concentration of 1X GE Blocking Agent and 1X Hi-RPM Hybridization Buffer, according to manufacturer’s instructions (miRNA Microarray System Protocol, Agilent Technologies). Arrays were washed according to manufacturer’s instructions (miRNA Microarray System Protocol, Agilent Technologies), dried out using a centrifuge at 1000 rpm for 2 min and scanned at 5µm resolution on an Agilent DNA Microarray Scanner (G2565BA, Agilent Technologies) equipped with extended dynamic range (XDR) software. Images provided by the scanner were analyzed using Agilent’s software Feature Extraction version 10.7.3.1. Data were quantile normalized as previously described [14].

Only miRNAs with an average intensity over the 20th percentile of the overall intensities and a detectable signal in at least 10 percent of the hybridized samples considered for further analysis. Batch effect was corrected using ComBat software [15].

### Consensus cluster

Consensus cluster using R v3.2.5 and ConsensusClusterPlus package was performed to establish subgroups [16]. Differential miRNAs expression patterns among groups was analyzed by Significance Analysis of Microarrays (SAM) with MeV 4.9 [17]. SAM performed a t-test correcting over permutations of the number of samples [18].

Targets of these differential miRNAs were searched in miRwalk database [19]. This information was used to perform a gene ontology analysis and to establish relationship with biological functions. Gene ontology analyses were done using Enrichr webtool developed by Ma’ayan lab [20].

### Statistical analyses

Statistical analyses were done using GraphPad Prism v6. All p-values were two-sided and considered statistically significant under 0.05.

## Results

### Patient cohort

164 renal tumor samples were studied. One hundred of these samples corresponded to clear-cell carcinomas (ccRCC), 16 to papillary tumors, and 21 chromophobe tumors. Subtype information was not available for 27 tumors. Of these 164 samples, clinical data were available for 142 patients. Twenty-three percent of the patients suffered a relapse and the median of follow-up was 54 months.

**Table 1:**
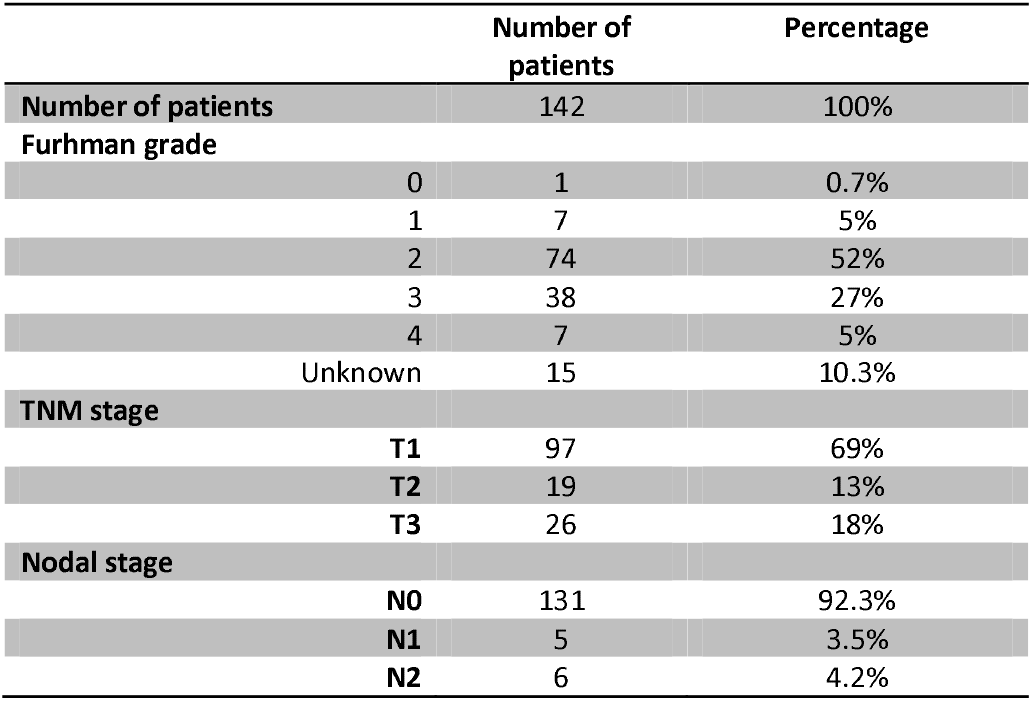
Patient characteristics.

### Characterization of differences between histological subtypes

A SAM was done to characterize different miRNA expression patterns between histological groups. It was not possible to find differential miRNAs between papillary and the two other histological subtypes. SAM showed that chromophobe tumors are a very homogeneous molecular group with a higher expression of miR 1229, miR 10a, miR 182, miR 1208, miR 222, miR 221, miR 891b, miR 629-5p and miR 221-5p (Fig 1).

**Fig 1:**
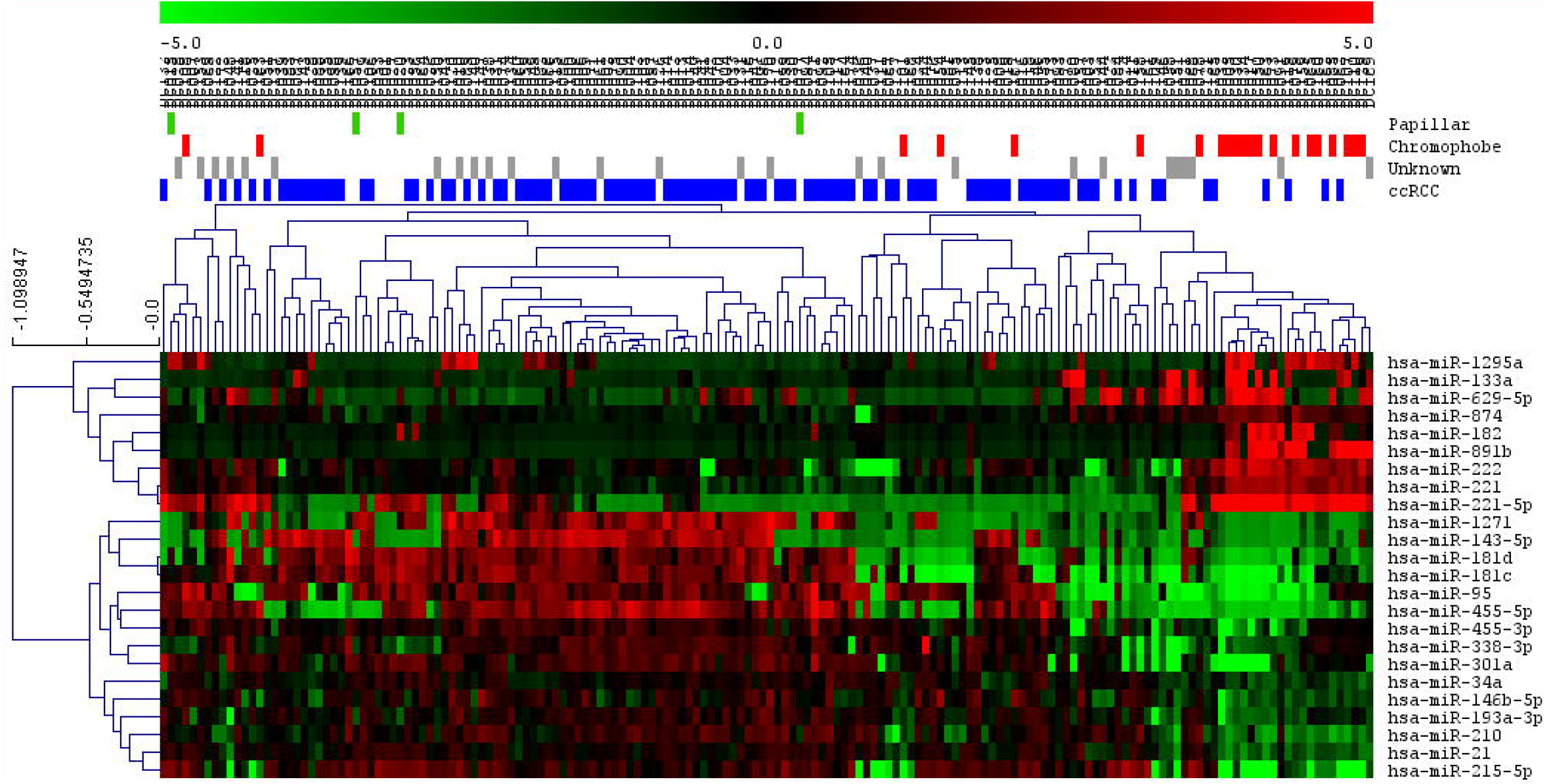
SAM of chromophobe subtype against the rest of tumors. ccRCC= Clear cell renal carcinoma.

On the other hand, ccRCC tumors were split into two different groups in the SAM graph, suggesting the existence of two molecular groups in ccRCC according the miRNA expression (Fig 2).

**Fig 2:**
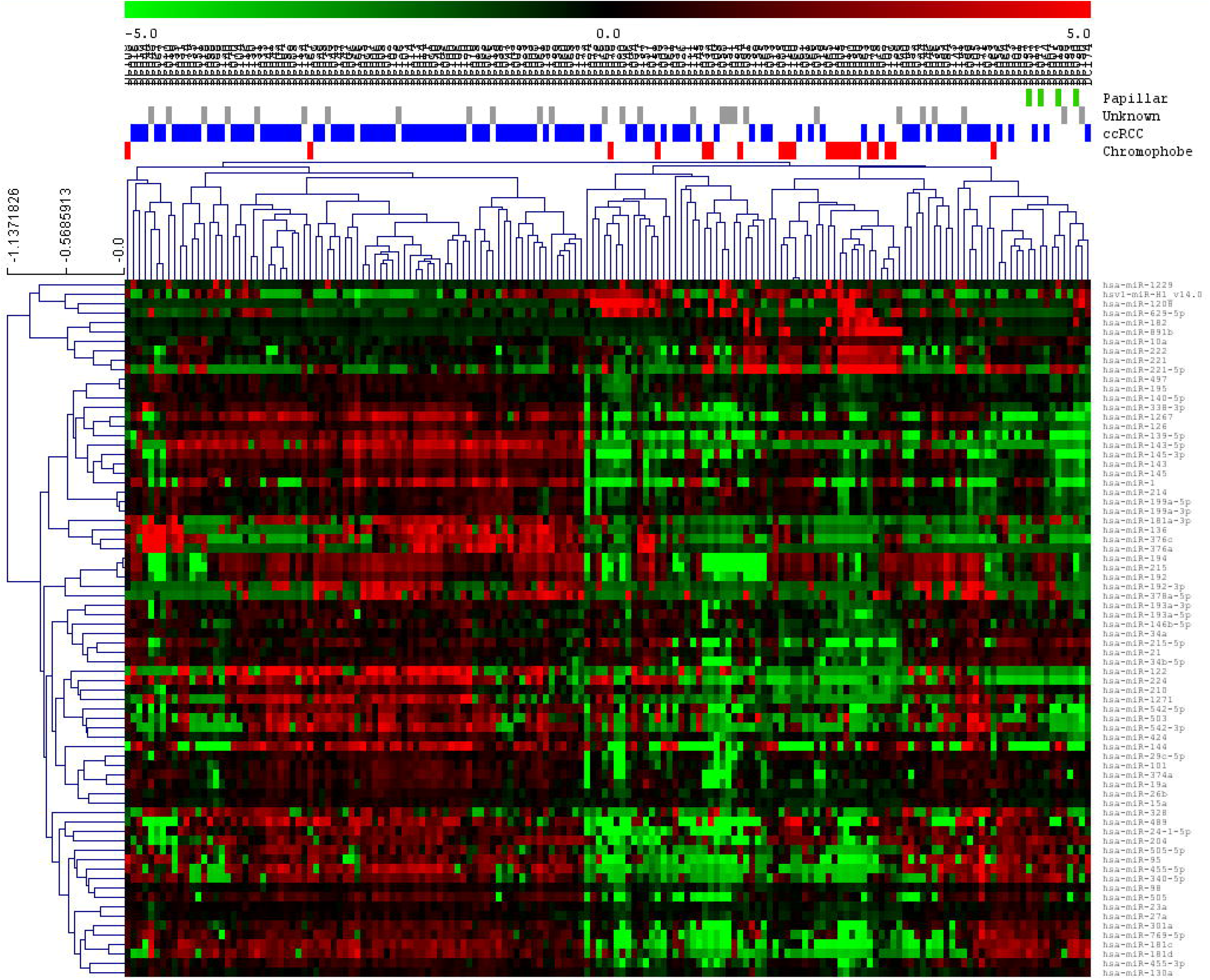
SAM of ccRCC tumors against the rest of them. ccRCC= Clear cell renal carcinoma.

### ccRCC groups’ characterization

With the aim of establishing these possible subgroups in ccRCC, a consensus cluster was done. Consensus cluster grouped patients by the similarity in their expression patterns and it allows the definition of the optimum number of groups, showing that two different molecular patient groups existed in ccRCC: ccRCC1 (44 patients) and ccRCC2 (56 patients) (Fig 3).

**Fig 3:**
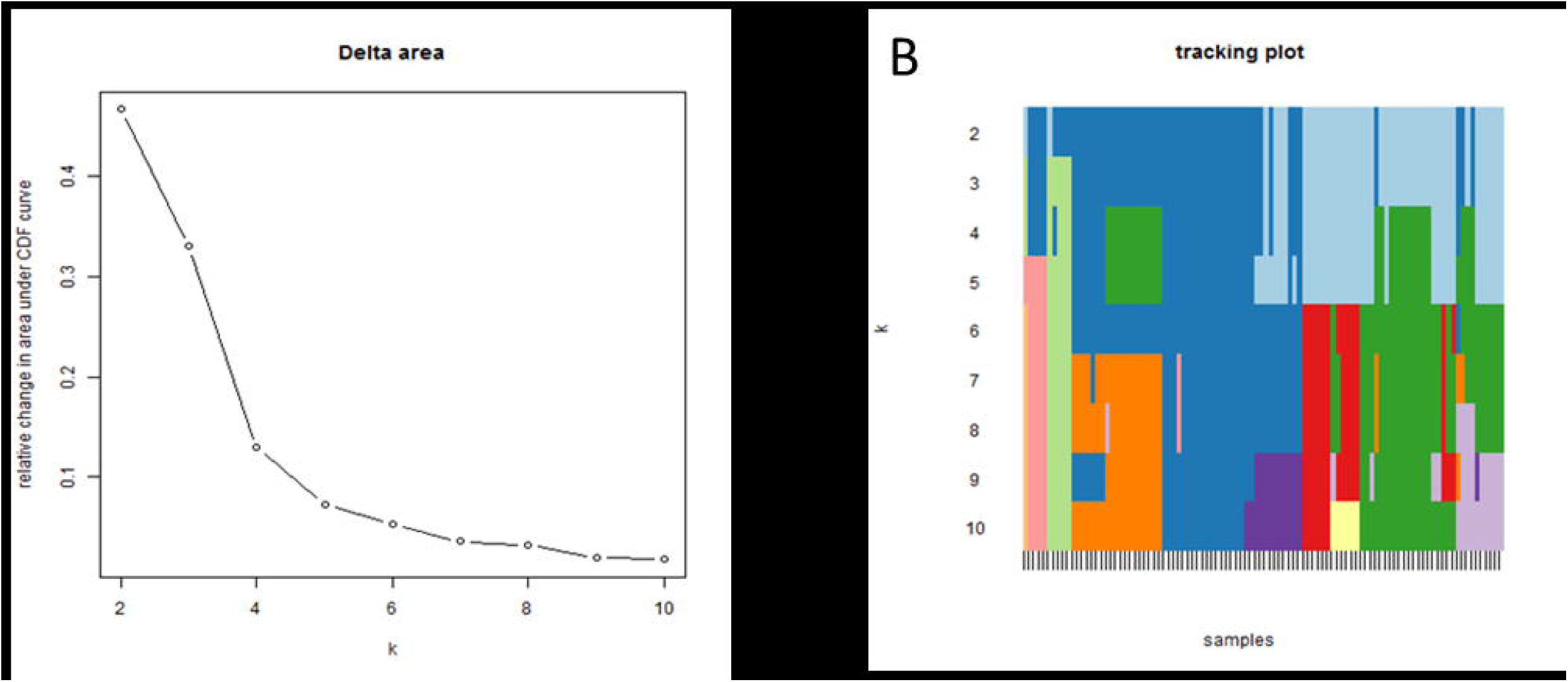
A. Delta graph suggested two groups as the optimum number of groups in these subtype. B. Tracking plot showed different sample classifications making different number of groups.

Contingency analyses showed that this new ccRCC classification was independent from clinical data, such as tumor size or nodal status; i.e, there are no differences in tumor size or nodal status betwween these two groups (p=0.55 and p=0.39 respectively). However, ccRCC2 tumors had a lower Furhman grade than ccRCC1 tumors (p=0.04).

Moreover, a SAM established 136 differentially expressed miRNAs between these two groups (Fig 4).

**Fig 4:**
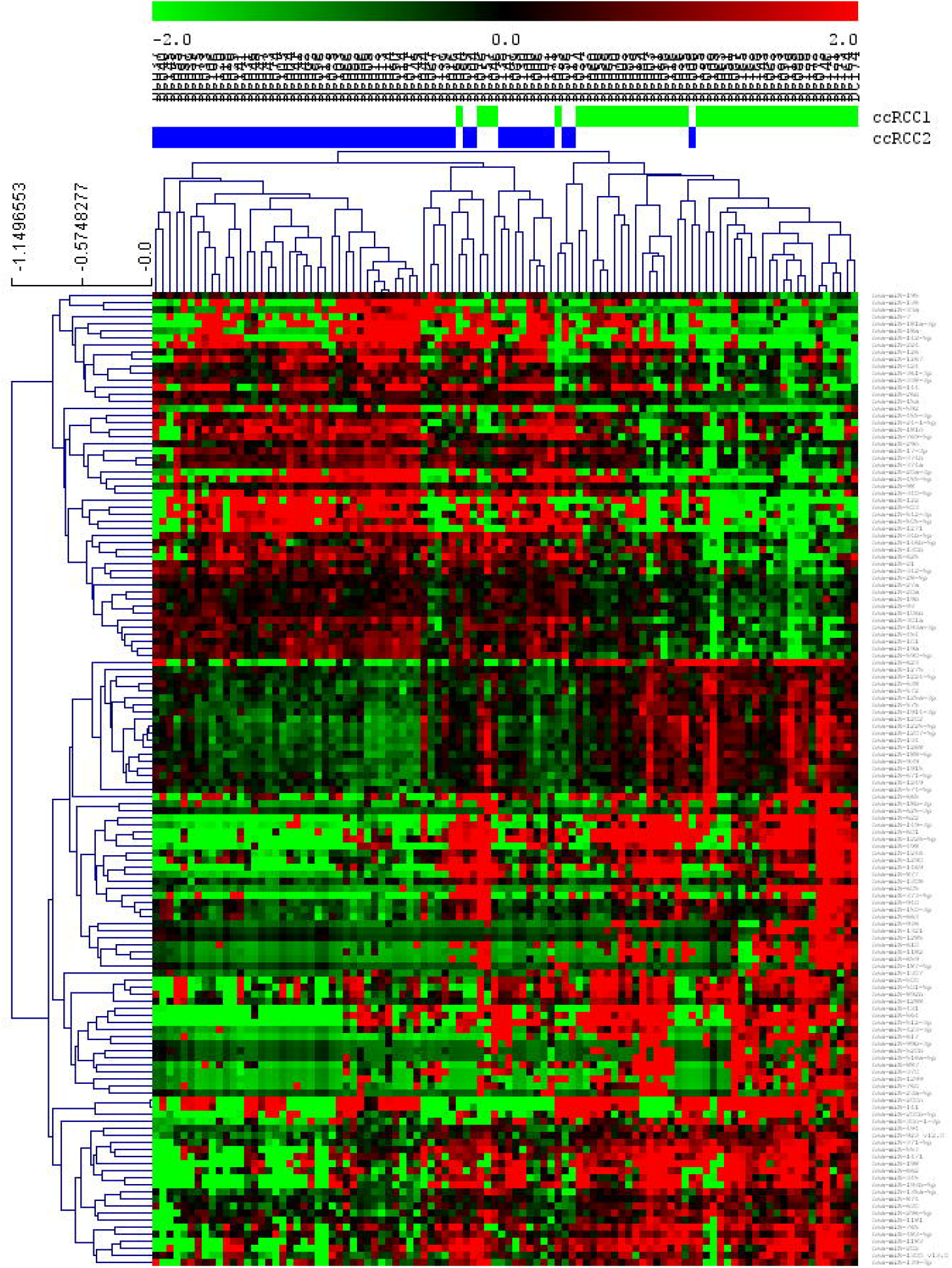
SAM between two ccRCC identified groups.

Experimentally validated targets of these 136 miRNAs were determined using miRwalk database, and a gene ontology analysis of these genes was performed afterwards. This analysis showed that these genes were mainly related with focal adhesion, transcription, apoptosis and angiogenesis processes (Fig 5).

**Fig 5:**
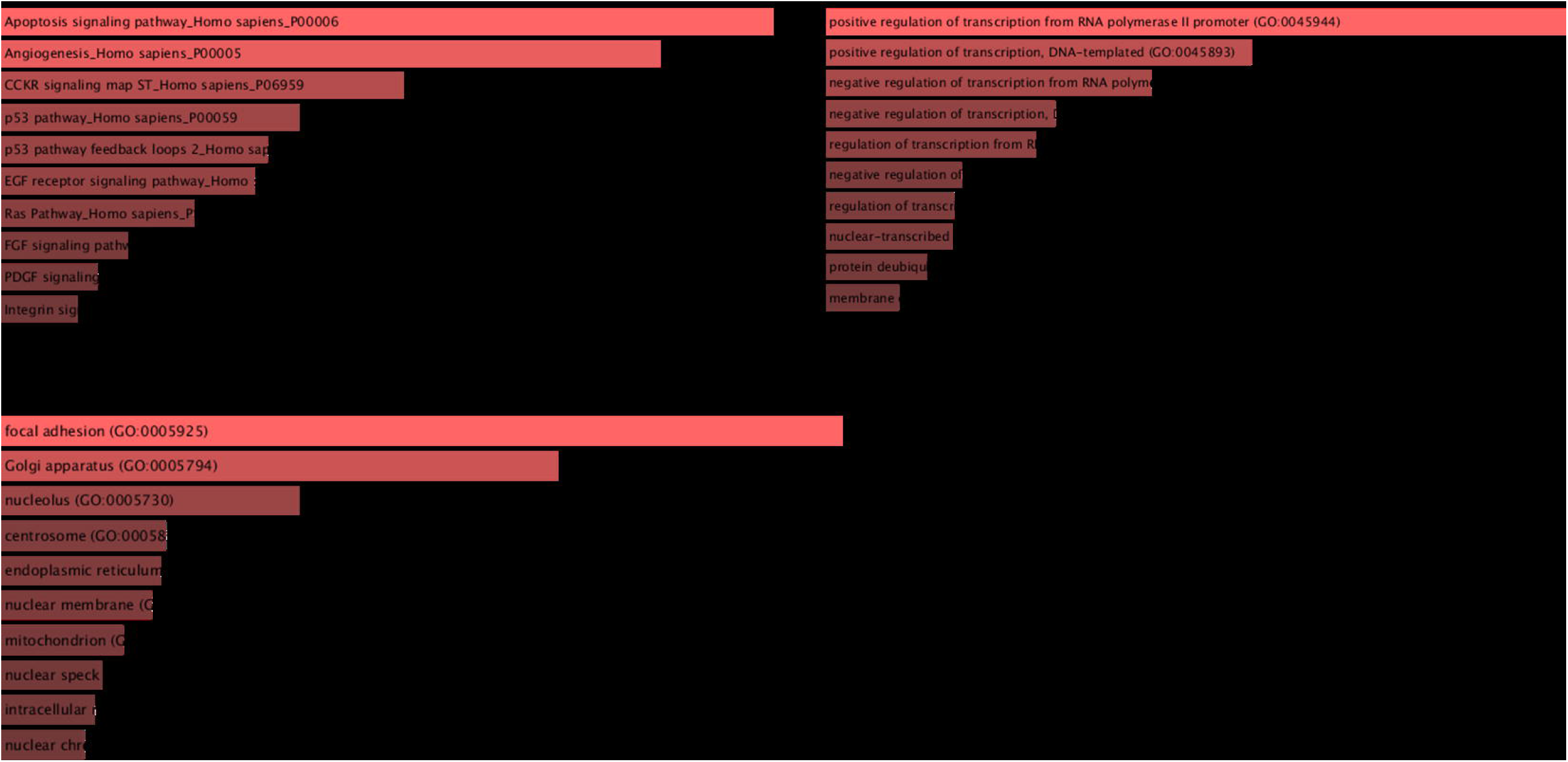
Gene ontology of gene targets of the 136 differential miRNAs.

Additionally, the two subgroups of ccRCC were associated with a different survival, although not statistically significant (Fig 6).

**Fig 6:**
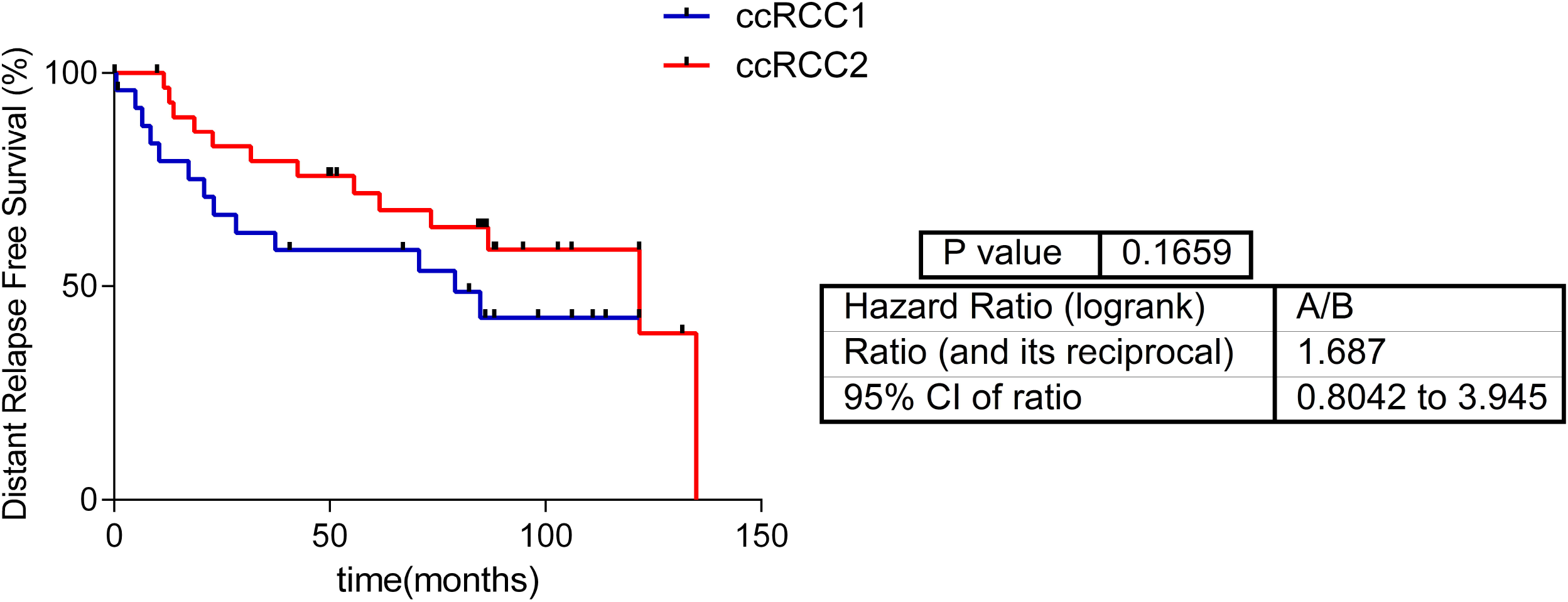
Survival curves of the two ccRCC groups defined by Consensus Cluster.

### New ccRCC groups had differential expression of miRNAs involved in angiogenesis

Interestingly, the two ccRCC subgroups presented differences in expression of some miRNAs previously associated with angiogenesis. In general, ccRCC2 had more expression of miR185, miR126 and miR130a, all of them proangiogenic miRNAs (Fig 7).

**Fig 7:**
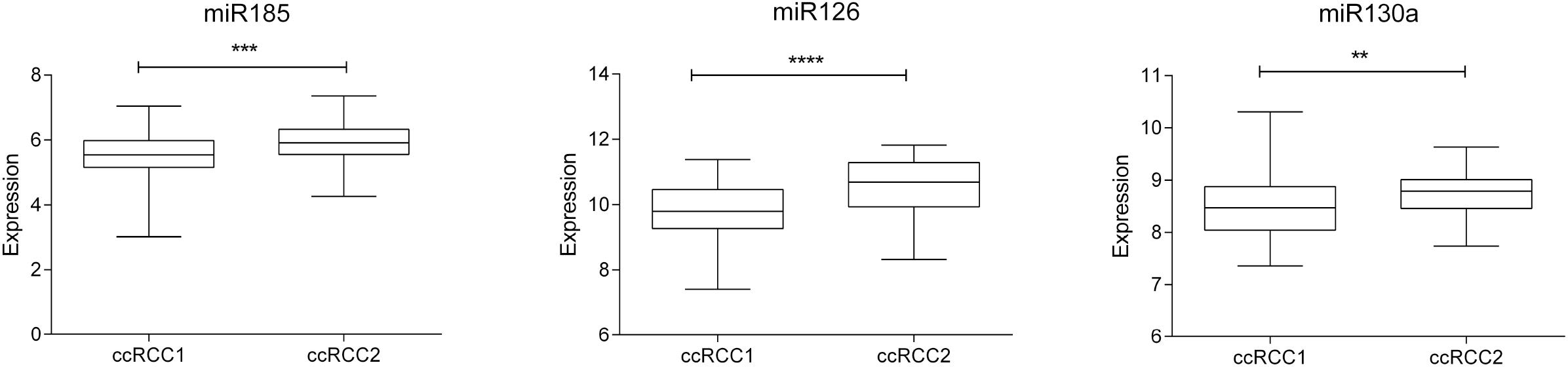
miRNAs related with angiogenesis differentialy expressed between two ccRCC groups

## Discussion

Renal-cell carcinoma comprises several histological subgroups [8]. The Cancer Genome Atlas analyzed these histological subtypes and characterized molecular differences between them [9, 10]. However, all these advances have not been translated into clinical applications yet. For this reason, further insight into the molecular biology of these tumors is still needed.

There are previous classifications of renal histological subtypes based on miRNA signatures, although a reduce number of miRNAs were used in these analyses [21-23]. In this study, 396 miRNAs were analyzed in 164 RC FFPE samples by microRNA arrays. The main advantage of the measurement of miRNAs is that they are more stable than longer RNAs or DNA in paraffin samples [12].

SAM showed differences at the miRNA expression level in chromophobe and ccRCC, but no in papillary tumors. The fact that it was not possible to define differential miRNAs in the papillary subgroup could be due to the reduced number of this type of tumors in our cohort and the intrinsic heterogeneity of this group.

miR10-a, miR222, and miR221 have been previously described as overexpressed in chromophobe subtype, what agrees with our results [22, 23].

Regarding ccRCC tumors, differential expression pattern analysis suggested the existence of two different groups inside this histological subtype. This was confirmed by Consensus Cluster, which defined two groups with different expression in miRNAs whose established targets are related to angiogenesis, apoptosis, transcription and focal adhesion.

Interestingly, three of the miRNAs (miR185, miR126, and miR130a) differentially expressed between our two ccRCC groups have been previously related with pro-angiogenesis processes.

Expression levels of miR185 have been correlated with tumor size, Fuhrman grade, and TNM staging. The overexpression of this miRNA inhibited proliferation and induced apoptosis [24]. Moreover, elevated miR185 levels were associated with high vascular endothelial growth factor receptor 2 (VEGFR) expression and therefore a pro-angiogenic activity in ccRCC [25].

On the other hand, miR126 inhibits the expression of vascular cell adhesion molecule 1 (VCAM1) implicated in leukocyte adherence to endothelial cells [26]. This miRNA has a pro-angiogenic function [27]. It is downregulated in metastatic ccRCC versus primary tumors. Its overexpression is negatively correlated with tumor size and is associated with longer distant relapse-free survival and overall survival. miR126 overexpression is also related with a reduction in cellular proliferation [28]. According to these facts, miR126 was underexpressed in our ccRCC1 group which had a worst prognosis.

miR130a were identified as proangiogenic miRNA due to its inhibitory effect in the anti-angiogenic homeobox GAX and HoxA5 [29]. A relationship between this miRNA and renal carcinoma had not been previously described.

Considering the controversial efficacy of antiangiogenic drugs in the adjuvant setting of renal-cell carcinoma [30], defining a proangiogenic group may be important to select patients more likely to benefit from these treatments. In the future, a class predictor could be developed to define this pro-angiogenic group. Such a predictor should be validated in an independent cohort.

In this work, we have characterized differences between RC histological subtypes using miRNAs and have defined two ccRCC groups with different expression of pro-angiogenic miRNAs. Differences between subtypes could be used as therapeutic targets or as a method to select patients for personalized treatments in the future.

## Data Availability

All relevant data are included as supplementary files.

## Supporting information

S1 Table: miRNAs measured in 164 renal carcinoma paraffin samples.

S2 Table: 136 differential miRNAs between ccRCC groups.

## References

1. Siegel RL, Miller KD, Jemal A. Cancer statistics, 2019. CA Cancer J Clin. 2019;69(1):7–34. Epub 2019/01/08. doi: 10.3322/caac.21551. PubMed PMID: 30620402.

2. Janzen NK, Kim HL, Figlin RA, Belldegrun AS. Surveillance after radical or partial nephrectomy for localized renal cell carcinoma and management of recurrent disease. Urol Clin North Am. 2003;30(4):843–52. PubMed PMID: 14680319.

3. Janowitz T, Welsh SJ, Zaki K, Mulders P, Eisen T. Adjuvant therapy in renal cell carcinoma-past, present, and future. Semin Oncol. 2013;40(4):482–91. doi: 10.1053/j.seminoncol.2013.05.004. PubMed PMID: 23972712; PubMed Central PMCID: PMCPMC3765962.

4. Staehler M, Motzer RJ, George DJ, Pandha HS, Donskov F, Escudier B, et al. Adjuvant sunitinib in patients with high-risk renal cell carcinoma: safety, therapy management, and patient-reported outcomes in the S-TRAC trial. Ann Oncol. 2018;29(10):2098–104. doi: 10.1093/annonc/mdy329. PubMed PMID: 30412222; PubMed Central PMCID: PMCPMC6247664.

5. Lawrence NJ, Martin A, Davis ID, Troon S, Sengupta S, Hovey E, et al. What Survival Benefits are Needed to Make Adjuvant Sorafenib Worthwhile After Resection of Intermediate-or High-Risk Renal Cell Carcinoma? Clinical Investigators’ Preferences in the SORCE Trial. Kidney Cancer. 2018;2(2):123–31. Epub 2018/08/01. doi: 10.3233/KCA-180038. PubMed PMID: 30740581; PubMed Central PMCID: PMCPMC6364092.

6. Motzer RJ, Haas NB, Donskov F, Gross-Goupil M, Varlamov S, Kopyltsov E, et al. Randomized Phase III Trial of Adjuvant Pazopanib Versus Placebo After Nephrectomy in Patients With Localized or Locally Advanced Renal Cell Carcinoma. J Clin Oncol. 2017;35(35):3916–23. Epub 2017/09/13. doi: 10.1200/JCO.2017.73.5324. PubMed PMID: 28902533; PubMed Central PMCID: PMCPMC6018511.

7. Gross-Goupil M, Kwon TG, Eto M, Ye D, Miyake H, Seo SI, et al. Axitinib versus placebo as an adjuvant treatment of renal cell carcinoma: results from the phase III, randomized ATLAS trial. Ann Oncol. 2018;29(12):2371–8. doi: 10.1093/annonc/mdy454. PubMed PMID: 30346481; PubMed Central PMCID: PMCPMC6311952.

8. Moch H, Cubilla AL, Humphrey PA, Reuter VE, Ulbright TM. The 2016 WHO Classification of Tumours of the Urinary System and Male Genital Organs-Part A: Renal, Penile, and Testicular Tumours. Eur Urol. 2016;70(1):93–105. Epub 2016/02/28. doi: 10.1016/j.eururo.2016.02.029. PubMed PMID: 26935559.

9. Ricketts CJ, De Cubas AA, Fan H, Smith CC, Lang M, Reznik E, et al. The Cancer Genome Atlas Comprehensive Molecular Characterization of Renal Cell Carcinoma. Cell Rep. 2018;23(1):313-26.e5. doi: 10.1016/j.celrep.2018.03.075. PubMed PMID: 29617669; PubMed Central PMCID: PMCPMC6075733.

10. Linehan WM, Spellman PT, Ricketts CJ, Creighton CJ, Fei SS, Davis C, et al. Comprehensive Molecular Characterization of Papillary Renal-Cell Carcinoma. N Engl J Med. 2016;374(2):135–45. Epub 2015/11/04. doi: 10.1056/NEJMoa1505917. PubMed PMID: 26536169; PubMed Central PMCID: PMCPMC4775252.

11. Wang WT, Chen YQ. Circulating miRNAs in cancer: from detection to therapy. J Hematol Oncol. 2014;7:86. Epub 2014/12/05. doi: 10.1186/s13045-014-0086-0. PubMed PMID: 25476853; PubMed Central PMCID: PMCPMC4269921.

12. Kakimoto Y, Tanaka M, Kamiguchi H, Ochiai E, Osawa M. MicroRNA Stability in FFPE Tissue Samples: Dependence on GC Content. PLoS One. 2016;11(9):e0163125. Epub 2016/09/20. doi: 10.1371/journal.pone.0163125. PubMed PMID: 27649415; PubMed Central PMCID: PMCPMC5029930.

13. Gámez-Pozo A, Berges-Soria J, Arevalillo JM, Nanni P, López-Vacas R, Navarro H, et al. Combined label-free quantitative proteomics and microRNA expression analysis of breast cancer unravel molecular differences with clinical implications. Cancer Res; 2015. p. 2243–53.

14. López-Romero P, González MA, Callejas S, Dopazo A, Irizarry RA. Processing of Agilent microRNA array data. BMC Res Notes. 2010;3:18. Epub 2010/01/22. doi: 10.1186/1756-0500-3-18. PubMed PMID: 20205787; PubMed Central PMCID: PMCPMC2823597.

15. Johnson WE, Li C, Rabinovic A. Adjusting batch effects in microarray expression data using empirical Bayes methods. Biostatistics. 2007;8(1):118–27. doi: 10.1093/biostatistics/kxj037. PubMed PMID: 16632515.

16. Monti S, Tamayo P, Mesirov J, Golub T. Consensus Clustering: A Resampling-Based Method for Class Discovery and Visualization of Gene Expression Microarray Data. Machine learning. 2003;52(1):91–118.

17. Saeed AI, Sharov V, White J, Li J, Liang W, Bhagabati N, et al. TM4: a free, open-source system for microarray data management and analysis. Biotechniques. 2003;34(2):374–8. PubMed PMID: 12613259.

18. Tusher VG, Tibshirani R, Chu G. Significance analysis of microarrays applied to the ionizing radiation response. Proc Natl Acad Sci U S A. 2001;98(9):5116–21. Epub 2001/04/17. doi: 10.1073/pnas.091062498. PubMed PMID: 11309499; PubMed Central PMCID: PMCPMC33173.

19. Dweep H, Sticht C, Pandey P, Gretz N. miRWalk--database: prediction of possible miRNA binding sites by “walking” the genes of three genomes. J Biomed Inform. 2011;44(5):839–47. Epub 2011/05/14. doi: 10.1016/j.jbi.2011.05.002. PubMed PMID: 21605702.

20. Chen EY, Tan CM, Kou Y, Duan Q, Wang Z, Meirelles GV, et al. Enrichr: interactive and collaborative HTML5 gene list enrichment analysis tool. BMC Bioinformatics. 2013;14:128. Epub 2013/04/15. doi: 10.1186/1471-2105-14-128. PubMed PMID: 23586463; PubMed Central PMCID: PMCPMC3637064.

21. Silva-Santos RM, Costa-Pinheiro P, Luis A, Antunes L, Lobo F, Oliveira J, et al. MicroRNA profile: a promising ancillary tool for accurate renal cell tumour diagnosis. Br J Cancer. 2013;109(10):2646–53. Epub 2013/10/15. doi: 10.1038/bjc.2013.552. PubMed PMID: 24129247; PubMed Central PMCID: PMCPMC3833202.

22. Youssef YM, White NM, Grigull J, Krizova A, Samy C, Mejia-Guerrero S, et al. Accurate molecular classification of kidney cancer subtypes using microRNA signature. Eur Urol. 2011;59(5):721–30. Epub 2011/01/13. doi: 10.1016/j.eururo.2011.01.004. PubMed PMID: 21272993.

23. Powers MP, Alvarez K, Kim HJ, Monzon FA. Molecular classification of adult renal epithelial neoplasms using microRNA expression and virtual karyotyping. Diagn Mol Pathol. 2011;20(2):63–70. doi: 10.1097/PDM.0b013e3181efe2a9. PubMed PMID: 21532496.

24. Ma X, Shen D, Li H, Zhang Y, Lv X, Huang Q, et al. MicroRNA-185 inhibits cell proliferation and induces cell apoptosis by targeting VEGFA directly in von Hippel-Lindau-inactivated clear cell renal cell carcinoma. Urol Oncol. 2015;33(4):169.e1-11. Epub 2015/02/17. doi: 10.1016/j.urolonc.2015.01.003. PubMed PMID: 25700976.

25. Yuan HX, Zhang JP, Kong WT, Liu YJ, Lin ZM, Wang WP, et al. Elevated microRNA-185 is associated with high vascular endothelial growth factor receptor 2 expression levels and high microvessel density in clear cell renal cell carcinoma. Tumour Biol. 2014;35(12):12757–63. Epub 2014/09/14. doi: 10.1007/s13277-014-2602-9. PubMed PMID: 25217984.

26. Harris TA, Yamakuchi M, Ferlito M, Mendell JT, Lowenstein CJ. MicroRNA-126 regulates endothelial expression of vascular cell adhesion molecule 1. Proc Natl Acad Sci U S A. 2008;105(5):1516–21. Epub 2008/01/28. doi: 10.1073/pnas.0707493105. PubMed PMID: 18227515; PubMed Central PMCID: PMCPMC2234176.

27. Fish JE, Santoro MM, Morton SU, Yu S, Yeh RF, Wythe JD, et al. miR-126 regulates angiogenic signaling and vascular integrity. Dev Cell. 2008;15(2):272–84. doi: 10.1016/j.devcel.2008.07.008. PubMed PMID: 18694566; PubMed Central PMCID: PMCPMC2604134.

28. Khella HW, Scorilas A, Mozes R, Mirham L, Lianidou E, Krylov SN, et al. Low expression of miR-126 is a prognostic marker for metastatic clear cell renal cell carcinoma. Am J Pathol. 2015;185(3):693–703. Epub 2015/01/05. doi: 10.1016/j.ajpath.2014.11.017. PubMed PMID: 25572155.

29. Chen Y, Gorski DH. Regulation of angiogenesis through a microRNA (miR-130a) that down-regulates antiangiogenic homeobox genes GAX and HOXA5. Blood. 2008;111(3):1217–26. Epub 2007/10/23. doi: 10.1182/blood-2007-07-104133. PubMed PMID: 17957028; PubMed Central PMCID: PMCPMC2214763.

30. Bex A. Adjuvant sunitinib in renal cell carcinoma: from evidence to recommendation. Ann Oncol. 2017;28(4):682–4. doi: 10.1093/annonc/mdx014. PubMed PMID: 28104620.

